# ONE CHAIN, REPEATED SPILLOVER, OR AN UNRELATED EARLY SIGNAL?

**DOI:** 10.64898/2026.08.02.26359503

**Authors:** Johan G.L. Verheyden, Celestin Nzanzu Mudogo, Wolfgang Jacquet

## Abstract

**Background:** A retrospective field investigation described substantial illness and mortality in and around Mongbwalu from January 2026, several months before Bundibugyo virus disease (BVD) was formally recognised in May. Historical BDBV evidence includes a genomic reconstruction compatible with multiple spillovers, while other orthoebolavirus outbreaks have involved survivor-associated resurgence, concurrent lineages and substantial syndromic misclassification. We evaluated whether the early Mongbwalu signal was most compatible with continuous acute transmission, a later introduction, survivor-mediated reseeding, or mixed/non-ancestral mortality.

**Methods:** We combined a structured rapid review of 35 sources with official confirmed surveillance, a published report of an unpublished retrospective investigation, and a soft genomic timing constraint. A resample-move approximate Bayesian computation sequential Monte Carlo model simulated 20 weekly periods across Mongbwalu, Bunia, Rwampara and Nizi. Four hypotheses were compared: continuous acute transmission (H1), repeated zoonotic introduction or temporarily elevated shared exposure (H2), survivor-mediated sexual transmission from persistent virus in semen (H3), and mixed or substantially non-ancestral retrospective mortality (H4). Three independent chains of 450 particles were run for each hypothesis over seven decreasing tolerances. Model recovery, prior sensitivity, posterior-predictive checks and leave-one-component-out analyses assessed identifiability and robustness.

**Results:** Under equal priors, H4 received 36.2%, H2 32.6%, H3 21.6% and H1 9.6%. Under the literature-neutral prior, H2 received 50.0%, H4 34.7%, H1 11.1% and H3 4.2%. H2 and H4 jointly accounted for 84.8% under the main prior and 68.7%-89.3% across all prior families, demonstrating a robust preference for a discontinuous origin history. The approximate evidence ratio was only 1.11:1 for H4 over H2, so H2’s lead under the main analysis reflected both close empirical fit and direct BDBV precedent incorporated into the prior. H1 was correctly recovered in 92% of equal-prior synthetic datasets, compared with 52% for H2, 42% for H3 and 24% for H4. Removing confirmed May counts eliminated H2’s clear advantage, identifying the rapid May expansion as the principal evidence for a later successful seed.

Interpretation: The findings favour a discontinuous origin: the January/February Mongbwalu signal was epidemiologically meaningful, but the lineage that expanded in May most probably arose from a later successful introduction or renewed primary exposure rather than from one uninterrupted acute chain. H2 is the preferred literature-informed explanation because it best reconciles the structured early signal,

March-centred sampled ancestry and rapid May growth, while direct BDBV precedent makes the mechanism credible. H4 remains a substantial alternative because the early events were not laboratory confirmed and H2 and H4 are only partly identifiable. H3 is temporally and biologically possible but lacks survivor-specific evidence, and H1 is not the leading explanation under any tested prior family.

## 1. INTRODUCTION

Reconstructing the origin of a filovirus outbreak is not equivalent to extrapolating the confirmed case curve backwards. Laboratory-confirmed surveillance usually begins after transmission has already occurred, whereas retrospective investigations preferentially recover severe illness, deaths, healthcare-worker infections and funeral-associated clusters. Those observations can establish that an unusual event probably occurred without determining whether the observed events shared direct ancestry with the viruses sampled later. Delayed recognition has been associated with larger and longer Ebola disease outbreaks, but a delay signal alone cannot establish one continuous lineage. [33]

Bundibugyo virus disease is particularly instructive because its early clinical presentation is not pathognomonic and broad suspected-case definitions are imperfect. During the 2007–2008 outbreak in Uganda, BDBV was identified as a distinct ebolavirus, but laboratory classification substantially narrowed the initially suspected population: 42 of 192 suspected cases were laboratory positive, 74 were probable and 77 were laboratory negative. [2–4] Retrospective symptom-based evidence must therefore be treated as graded compatibility evidence rather than as a confirmed epidemic curve.

The 2012 Isiro outbreak in the Democratic Republic of the Congo provides the most direct historical precedent for the present question. Expanded genomic sequencing challenged a simple single-introduction interpretation and was compatible with multiple BDBV spillovers; a patient record also moved the inferred emergence substantially earlier than the initially accepted chronology. [1] Household and healthcare transmission were important, and women represented 85.3% of community cases in that small, context-specific outbreak. [5] These observations show that BDBV origin histories can be more complex than a single index case and that retrospective chronology can change when additional records and genomes are incorporated.

Evidence from other orthoebolaviruses reinforces this point. A review of 35 Ebola disease outbreaks found wildlife spillover to be the usual origin but identified survivor-associated resurgence as a substantial minority mechanism. [7] In the 2020 Équateur Province outbreak, genomic analysis identified two concurrent variants: one consistent with a new zoonotic introduction and another compatible with survivor persistence. [8] The 2021 Guinea resurgence was genetically linked to the 2013–2016 epidemic and accumulated far less divergence than sustained transmission would have produced. [9] Multiple human infection events have also been documented during central African wildlife epizootics. [10]

Mongbwalu is a mining and mobility centre, making temporally clustered primary exposure a plausible mechanism to test. The prolonged Marburg virus outbreak around Durba and Watsa illustrates how repeated primary introductions among miners can sustain what otherwise appears to be one extended epidemic. [11–13] This is a structural analogy rather than a numerical prior. BDBV has not been detected in a confirmed reservoir, Egyptian rousette bats are not a supported source of ebolavirus spillover, and current evidence does not justify assigning a particular bat species, mine reservoir or animal contact to the 2026 event. [7,14–17]

Survivor-mediated reseeding is biologically credible but quantitatively uncertain. Genomic evidence has linked EBOV transmission to a male survivor approximately six months after illness, and seminal RNA can persist for many months. [22–26] However, RNA detection is not equivalent to viable infectious virus, and some survivor cohorts reported no known sexual transmissions despite RNA-positive semen and periods of condomless sex. [27] No BDBV-specific persistence curve or per-exposure transmission probability was identified.

Gendered exposure affects both ordinary household transmission and the survivor-bridge hypothesis. Direct care, contact with wet symptoms, death in the household and close family relationships increase infection risk. [30–31] The literature does not establish general female biological susceptibility; female overrepresentation is better explained by caregiving, household and occupational exposure. [32] WHO reported that more than 60% of early suspected cases in the 2026 outbreak were female. [20] Sex composition was therefore treated as part of the exposure structure rather than as intrinsic vulnerability.

The 2026 evidence consists of three layers with different inferential roles. First, a retrospective investigation reported through ScienceInsider described more than 500 suspected events from mid-January to 15 May after 97 interviews and review of documentary material, including dated household and funeral-associated deaths and illness among frontline workers. [21] The underlying weekly line list and laboratory status of the January events were not public. Second, genomic analyses placed the sampled common ancestor in early to mid-March, with uncertainty extending into February and April, while only a small number of available genomes came from Mongbwalu. [18–19] Third, official surveillance recorded rapid confirmed growth after recognition. [20,35–37]

These layers answer different questions. The retrospective material informs whether an unusual early signal existed; genomic timing informs the ancestry of sampled viruses; and confirmed surveillance constrains the dynamics of the recognised epidemic. We therefore did not ask whether every early event was BVD. We asked which origin pathway best reconciled the three evidence layers and whether the available summaries could distinguish those pathways when the true mechanism was known.

## 2. METHODS

### 2.1 Study design and estimand

We conducted a comparative outbreak-origin analysis using public, aggregated and secondary evidence. The estimand was the relative support, conditional on the stated hypothesis set, prior family, simulator and summary statistics, for four mechanisms linking or separating the January/February Mongbwalu signal and the outbreak recognised in May. The estimand was not the probability that any named retrospective event was BVD and was not the probability that all reported early deaths belonged to the confirmed outbreak.

### 2.2 Evidence hierarchy and observation status

Evidence was classified before modelling to prevent unconfirmed retrospective information from acquiring the same status as laboratory-confirmed surveillance. Official confirmed counts were treated as the strongest observational layer; genomic timing was treated as a model-derived ancestry constraint; dated household and funeral events were treated as unconfirmed temporal anchors; and the reported retrospective aggregate was treated as an uncertain compatibility signal. The confirmed series was drawn from the BDBV2026-Data CORE repository through 20 July 2026, including national and health-zone cumulative counts. [37] The independently documented growth analysis provided a corrected interpretation of the early confirmed series. [36] The evidence components are summarised in Table 1.

**Table 1.**
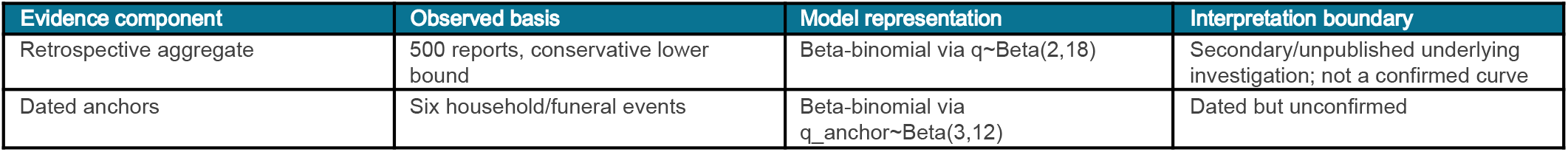
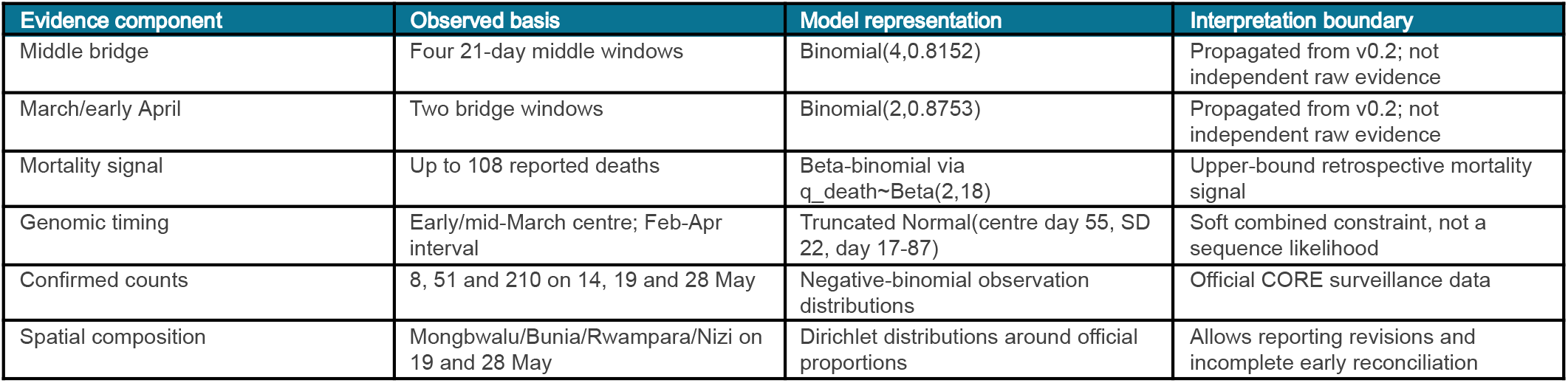
Evidence components, observed basis, probabilistic representation and interpretation boundaries.

**Figure 1.**
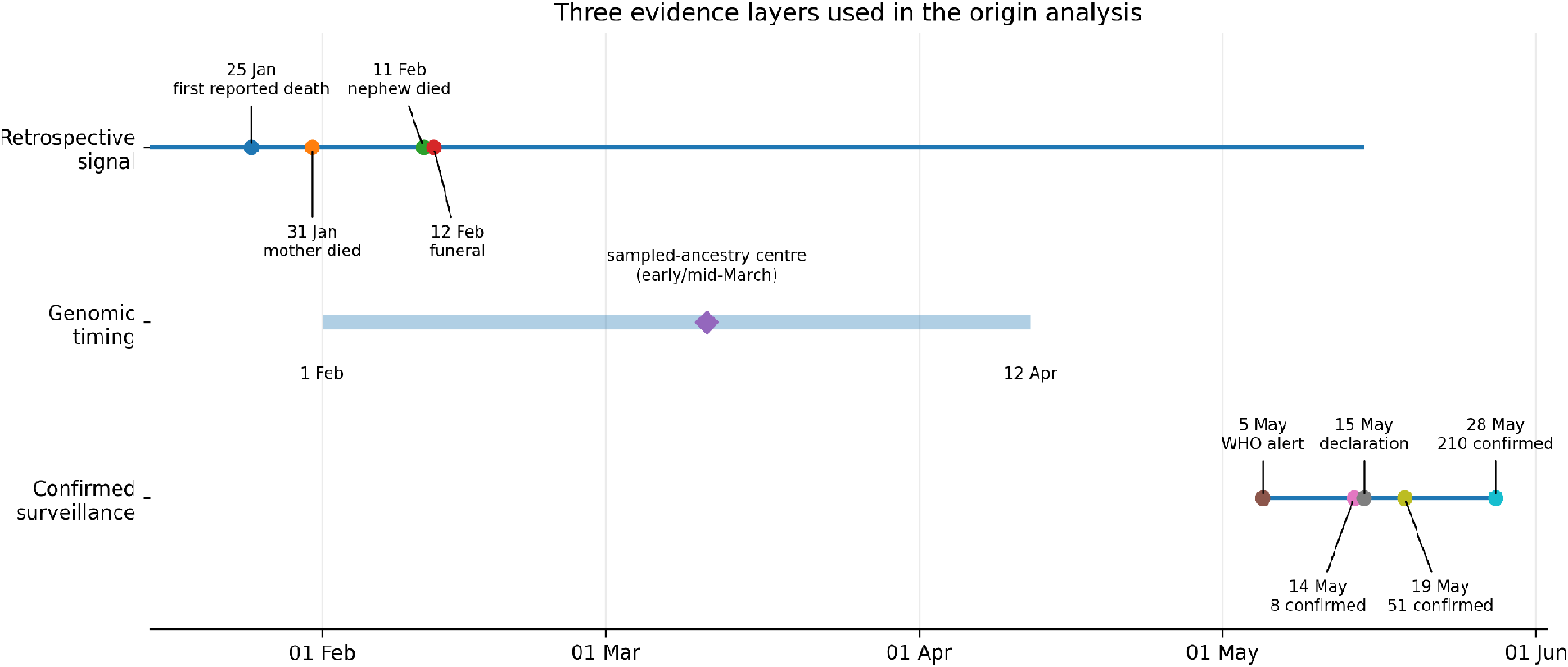
Timeline and evidence hierarchy. The January–May retrospective period, the genomic timing interval and confirmed surveillance are shown as separate evidence layers; the shaded regions do not represent continuous confirmed transmission.

### 2.3 Structured rapid literature review and prior development

A structured rapid review was completed on 2 August 2026. PubMed/MEDLINE, PubMed Central, WHO, CDC/Emerging Infectious Diseases, journal publisher sites, Virological.org and backward/forward citation chains were searched. Search concepts combined BDBV with origin, spillover, introduction, genomic and Isiro terms; Ebola or filovirus with multiple introduction, resurgence, persistence and sexual transmission; and BVD/EVD with women, caregiving, household transmission, suspected-case classification and delayed recognition.

Primary outbreak investigations, genomic studies, survivor cohorts, household-transmission studies, relevant experimental reservoir studies and official current-outbreak sources were retained. BDBV-specific primary evidence received the highest applicability grade. Other orthoebolavirus evidence was used where a biological mechanism lacked BDBV-specific estimates, while Marburg virus evidence was used only as a structural analogue for repeated primary exposure in a mining setting. Thirty-five sources were included: ten BDBV-specific primary sources, seventeen other-ebolavirus primary studies or syntheses, three related-filovirus analogues and five official/current or secondary sources.

The review was not registered, and screening and extraction were not independently duplicated. It was used to determine which mechanisms deserved explicit representation and to construct transparent sensitivity priors rather than to pool a common effect. The most consequential finding was direct BDBV precedent for a multiple-introduction history in Isiro. The review did not identify a defensible numerical frequency for two BDBV introductions in one locality over a three-month period.

### 2.4 Competing hypotheses and interpretation of Figure 2

The four hypotheses were defined as generative histories rather than narrative labels. All four were required to reproduce the rapid confirmed epidemic in May. They differed in what generated the January/February signal, whether an epidemiological bridge existed, and which event gave rise to the lineage sampled during the recognised outbreak. Their defining assumptions are summarised in Table 2.

**Table 2.**
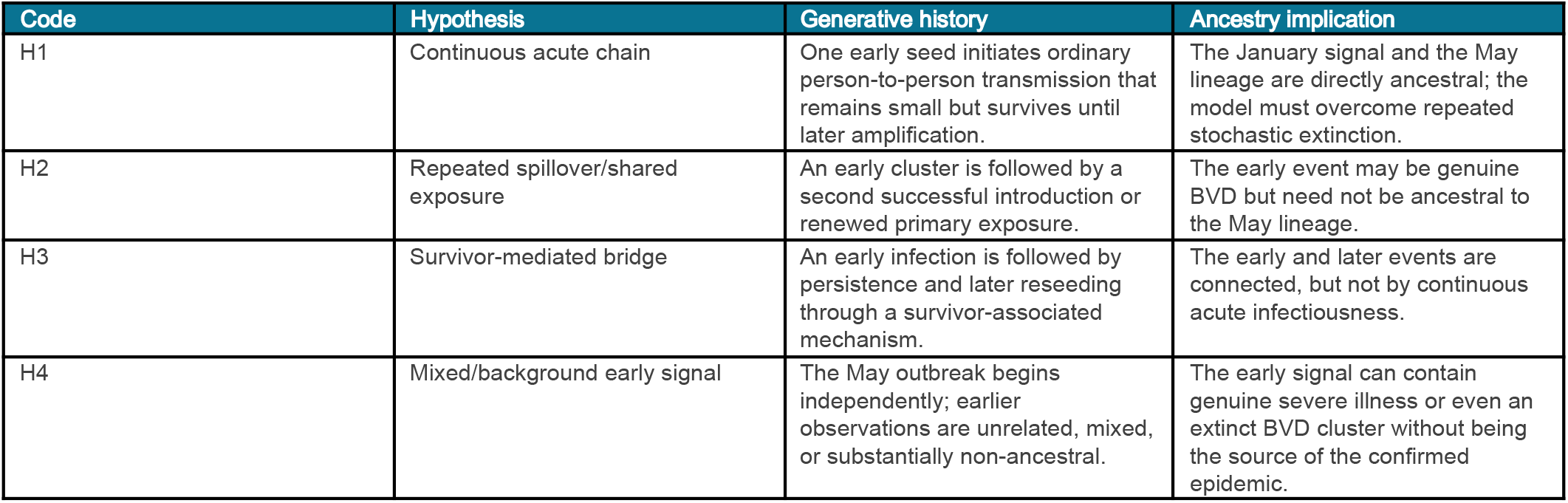
Competing origin hypotheses and their ancestry implications.

Figure 2 translates these definitions into weekly histories. In H1, a single line extends from the early seed to the May epidemic: the line is intentionally thin during the bridge period because the chain must remain small enough to avoid recognition. In H2, the early and later lines are separated, representing two introductions or two primary-exposure episodes; the first can terminate without affecting the second. H3 also contains two visible phases, but the arrow between them represents persistence in a survivor rather than a new external introduction. H4 shows scattered early observations without a transmission line leading into May, followed by an independent later outbreak. The figure therefore depicts ancestry assumptions, not estimated case counts.

H2 and H4 were deliberately separated even though both allow a later successful seed. H2 assigns substantive disease relevance to the early cluster, whereas H4 permits most of the early signal to arise from background or mixed causes. This distinction is central to the model-recovery analysis because the available observations may be insufficient to tell those histories apart.

**Figure 2.**
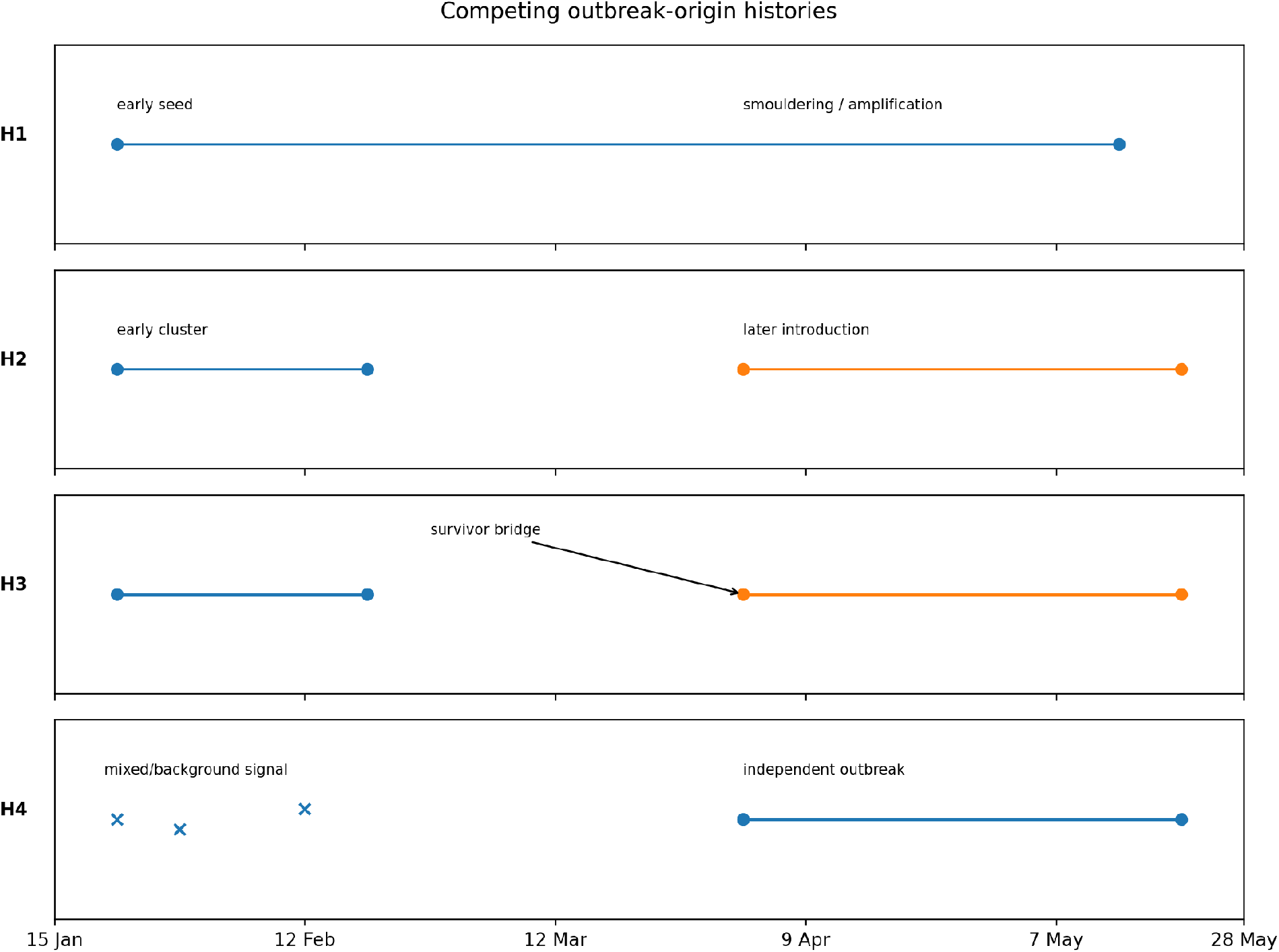
Competing origin histories. H1 requires one uninterrupted acute transmission lineage; H2 contains an early cluster and later introduction; H3 links early and later transmission through survivor persistence; H4 assigns the early signal to mixed or non-ancestral events and starts the recognised epidemic independently. The lines represent ancestry mechanisms, not reconstructed epidemic curves.

### 2.5 Weekly spatial transmission simulator

The simulator covered 20 weekly periods from 15 January through early June 2026 and four health zones in the early outbreak corridor: Mongbwalu, Bunia, Rwampara and Nizi. A weekly time step was selected because the retrospective evidence did not support daily precision, whereas monthly periods would obscure the January funeral-linked signal, the March–April bridge and the rapid May expansion.

Let I(t,z) denote incident infections in week t and health zone z. Conditional on the weekly reproduction parameter R(t) and overdispersion k, offspring were generated as:

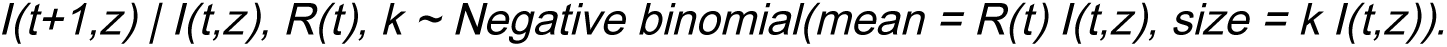

A movement parameter allocated a proportion of weekly offspring among the four zones using a stylised corridor matrix. R(t) was allowed to change from a lower pre-amplification value to a higher post-amplification value. The negative-binomial formulation allowed strong individual-level heterogeneity and a high probability that small chains would become extinct.

H1 contained one early introduction and a later amplification week. H2 contained early and later introductions. H3 contained an early introduction and a later survivor-associated seed. H4 contained only the later outbreak seed, with the early signal generated through the background observation process. Each model therefore had enough flexibility to reproduce the May outbreak but was required to do so through its stated mechanism.

### 2.6 Retrospective and prospective observation processes

Latent infections were separated from the events observed retrospectively or reported prospectively. Deaths were sampled from infections using an uncertain fatality fraction. Retrospective compatible observations combined disease-associated and background components:

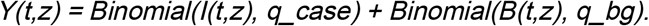

B(t,z) represents background severe illness or mortality, q_case the probability that a simulated infection contributes to the retrospective compatibility curve, and q_bg the corresponding probability for a background event. These are observation-process parameters. They do not represent assay sensitivity, case-definition specificity or an individual diagnostic probability.

Prospective confirmed reports were generated from infections through an uncertain reporting fraction. National cumulative counts on 14, 19 and 28 May were represented with negative-binomial observation uncertainty. The spatial distributions across Mongbwalu, Bunia, Rwampara and Nizi on 19 and 28 May were represented by Dirichlet distributions centred on official proportions. This retained information on early growth and geography while allowing for reporting delay, revision and incomplete reconciliation.

### 2.7 Genomic timing

The genomic component was a soft sampled-ancestry timing constraint rather than a sequence likelihood. The evidence distribution was centred in early to mid-March and truncated to the reported February–April interval. [19] H2 and H4 naturally generated a later sampled ancestor because the lineage responsible for the May epidemic was introduced later. H1 could generate a later sampled ancestor through loss of early branches, incomplete sampling and a lineage bottleneck. H3 generally shifted sampled ancestry later because reseeding followed a period of persistence.

A March-centred tMRCA therefore did not constitute evidence that no January event occurred. It constrained the ancestry of viruses actually sampled during the recognised outbreak. An earlier cluster could have become extinct, remained unsampled, or been replaced by a later lineage.

### 2.8 ABC-SMC algorithm

The retrospective observation process and weekly transmission histories did not yield a tractable exact likelihood. We therefore used approximate Bayesian computation with sequential Monte Carlo, an established likelihood-free approach for stochastic dynamical systems. [39–40]

For each hypothesis, model parameters were sampled, a weekly spatial history was generated, and the resulting summary vector was compared with draws from the uncertain evidence distribution. The comparison used the Gaussian ABC kernel:

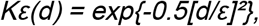

where d was a weighted standardised discrepancy across the retrospective compatible total, dated anchors, bridge-window occupancy, compatible deaths, genomic timing, national May counts and early spatial composition. Rather than compare simulations with one fixed pseudo-dataset, the kernel was integrated across 48 draws from the evidence distributions.

Three independent chains of 450 particles were run for each hypothesis. The tolerance schedule was [3.0, 2.4, 1.9, 1.5, 1.25, 1.05, 0.9]. At each stage, particles were reweighted, systematically resampled and rejuvenated by reflected random-walk Metropolis proposals. Approximate model evidence was accumulated sequentially, and model probabilities were calculated from the product of the model prior and approximate model evidence.

In practical terms, the algorithm repeatedly generated plausible versions of each origin history and gave greater weight to histories that could jointly reproduce the early retrospective pattern, the genomic timing and the rapid confirmed expansion in May.

### 2.9 Prior families

**Table 3.** Model-prior families used in the sensitivity analysis.

| Prior family | H1 | H2 | H3 | H4 | Purpose |
| --- | --- | --- | --- | --- | --- |
| Equal | 25% | 25% | 25% | 25% | Diagnostic family showing the evidence contribution without unequal model priors. |
| Continuity-favouring | 45% | 25% | 5% | 25% | Deliberately gives H1 the greatest prior probability. |
| Literature-neutral | 30% | 40% | 5% | 25% | Main family; recognises direct BDBV precedent for multiple introduction while retaining substantial H4 probability. |
| H2-permissive | 20% | 55% | 5% | 20% | Sensitivity family representing a strong temporary shared-exposure episode. |

Within-model priors were broad. Pre-amplification reproduction could be below or modestly above one; post-amplification reproduction could support rapid growth; overdispersion allowed frequent extinction; and observation parameters allowed substantial under-detection. H3 received low model prior probability in the substantive families because persistence evidence establishes feasibility but does not provide a BDBV-specific probability of successful survivor-to-partner transmission and onward establishment. The model-prior families are listed in Table 3.

### 2.10 Validation, model recovery and evidence ablation

Simulation-based inference requires validation of both the computational implementation and the ability of the chosen summaries to distinguish models. Simulation-based calibration and model-recovery exercises are recommended because apparently precise posterior results can arise from coding errors, weak identification or structurally overlapping models. [44–45]

We generated 50 independent synthetic datasets under each hypothesis. Each synthetic dataset was classified against an independent reference table containing 800 simulations per model. Recovery was evaluated under equal and literature-neutral priors. The confusion matrix records P(selected model | true generating model); a diagonal cell therefore measures the frequency with which the correct model was recovered.

The recovery analysis was not used to adjust the reported model probabilities mechanically. It was used to determine how strongly those probabilities could be interpreted. High recovery would support mechanism-specific interpretation, whereas confusion between H2 and H4 would imply that the evidence identifies a broader class of discontinuous histories more reliably than the precise mechanism.

Evidence ablation recalculated reference-table model support after removing the retrospective aggregate, dated anchors, middle bridge windows, March–April windows, deaths, genomic timing, national May counts, or either spatial composition. Additional scenarios down-weighted the entire retrospective bundle or the genomic component. Posterior-predictive histories were also generated from accepted particles and compared with the evidence summaries, following the general logic of posterior-predictive model assessment. [43]

### 2.11 Statistical interpretation and limits of ABC model probabilities

For hypothesis H, the final ABC model probability was calculated as:

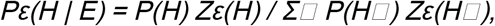

where P(H) is the model prior and Z_epsilon(H) the approximate evidence accumulated by the ABC-SMC chain. Posterior odds between two hypotheses therefore equal prior odds multiplied by an approximate evidence ratio. Reporting both quantities is essential because a model can lead under a literature-informed prior even when another model has slightly greater equal-prior evidence.

ABC model selection is sensitive to whether the summary statistics retain information that discriminates among models. Insufficient summaries can yield model probabilities that do not converge to the probabilities based on the full data. [41–42] We therefore avoided interpreting the percentages as direct probabilities that a specific zoonotic or survivor event occurred. We report the equal-prior comparison, the literature-informed prior sensitivity, recovery performance, chain stability and ablation results together.

The most defensible inferential level was determined empirically. If H2 and H4 were difficult to recover separately but both consistently exceeded H1, the robust conclusion was that a discontinuous history was favoured, not that one specific discontinuous mechanism had been proven. This class-level interpretation was specified before drafting the final conclusion.

Stated directly, a high model probability was treated as evidence that one history fitted the available summaries better than its competitors, not as proof that the corresponding unobserved event had occurred.

## 3. RESULTS

### 3.1 Monte Carlo performance and tolerance path

Final effective-sample fractions ranged from 87.5% for H1 to 95.2% for H4 before resampling. Mean final-stage MCMC acceptance ranged from 13.6% for H1 to 23.9% for H4. Between-chain standard deviations of log approximate evidence were 0.101 for H1, 0.069 for H2, 0.071 for H3 and 0.019 for H4. These differences were small relative to the evidence differences between H1 and the discontinuous models. Full final-stage diagnostics are reported in Table 4.

**Table 4.** Final-stage ABC-SMC chain diagnostics.

| Hypothesis | Mean final ESS fraction | Mean MCMC acceptance | Mean log ABC evidence | Between-chain SD |
| --- | --- | --- | --- | --- |
| H1 | 87.5% | 13.6% | -4.490 | 0.101 |
| H2 | 94.7% | 23.2% | -3.270 | 0.069 |
| H3 | 93.3% | 20.1% | -3.678 | 0.071 |
| H4 | 95.2% | 23.9% | -3.162 | 0.019 |

**Figure 3.**
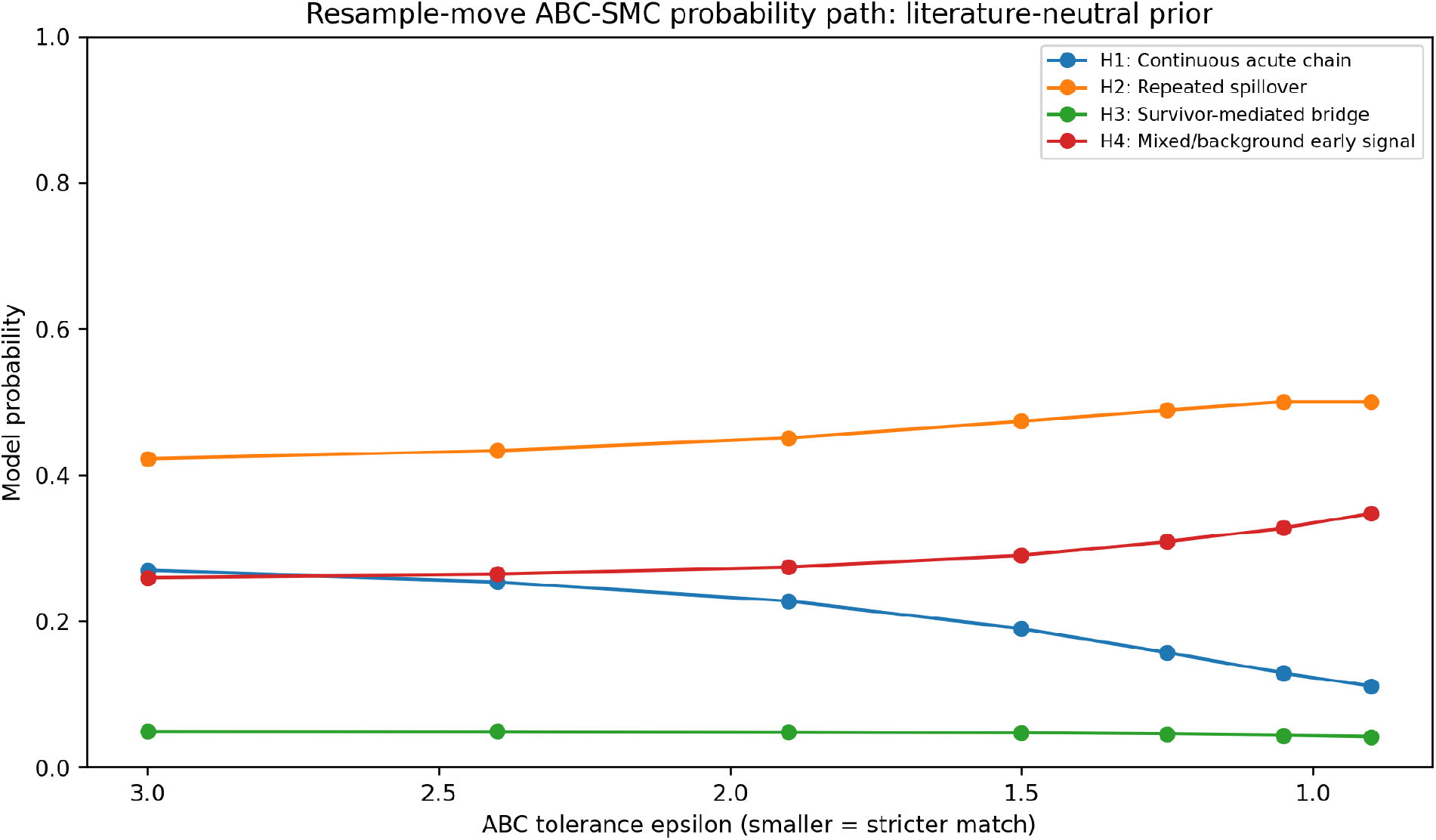
Model probability as the ABC tolerance decreased under the literature-neutral prior. H2 increased from 42.2% at epsilon=3.0 to 50.0% at epsilon=0.90, while H1 decreased from 27.0% to 11.1%. The ordering at the final stages was therefore not created by one isolated tolerance choice.

### 3.2 Model evidence, priors and hypothesis probabilities

Under equal priors, H4 received the highest probability (36.2%), followed by H2 (32.6%), H3 (21.6%) and H1 (9.6%). Relative to H1, the approximate evidence ratios were 3.76:1 for H4, 3.38:1 for H2 and 2.25:1 for H3. The underlying approximate evidence and prior-to-posterior movement are reported in Table 5.

**Table 5.**
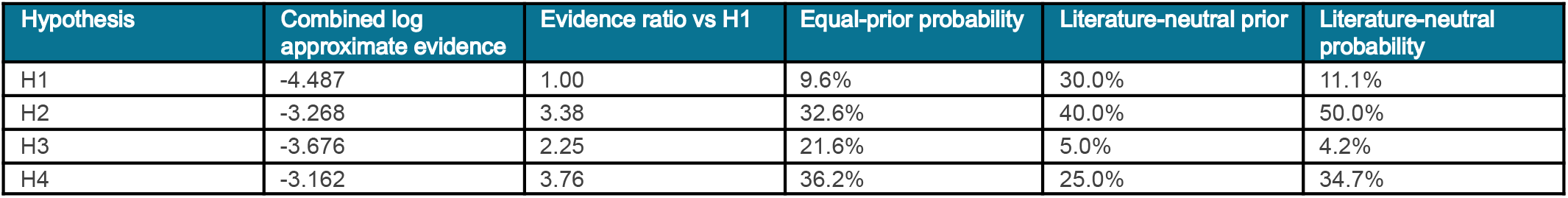
Approximate model evidence and model probabilities under equal and literature-neutral priors.

| Hypothesis | Combined log approximate evidence | Evidence ratio vs H1 | Equal-prior probability | Literature-neutral prior | Literature-neutral probability |
| --- | --- | --- | --- | --- | --- |
| H1 | -4.487 | 1.00 | 9.6% | 30.0% | 11.1% |
| H2 | -3.268 | 3.38 | 32.6% | 40.0% | 50.0% |
| H3 | -3.676 | 2.25 | 21.6% | 5.0% | 4.2% |
| H4 | -3.162 | 3.76 | 36.2% | 25.0% | 34.7% |

H4’s approximate evidence was only 1.11 times that of H2. The two models were therefore nearly tied on the summary-level evidence. Under the literature-neutral prior, the prior odds for H2 versus H4 were 1.60:1; multiplying by the approximate evidence ratio of 0.90:1 for H2 versus H4 yielded posterior odds of approximately 1.44:1 and probabilities of 50.0% and 34.7%. H2’s lead under the main analysis consequently reflected both direct BDBV precedent incorporated in the prior and a data fit close to H4, rather than an overwhelming evidence advantage.

#### Prior-family sensitivity and the robust class-level result

**Figure 4.**
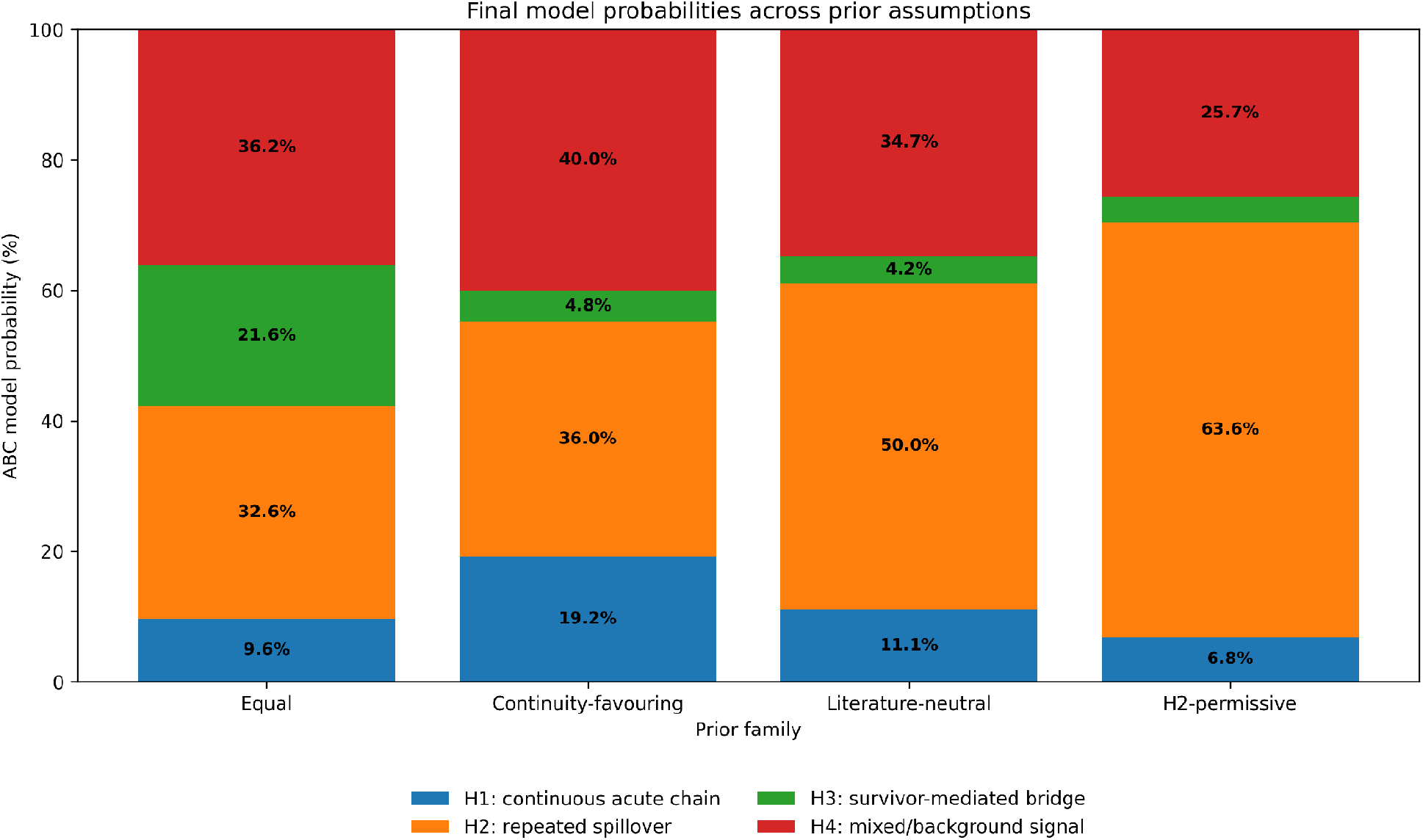
Final model probabilities across prior assumptions. Percentages are shown directly within the stacked bars. The legend is placed below the plotting area. Although the relative ordering of H2 and H4 changes with the prior, their combined support remains dominant.

The probability assigned to the combined discontinuous class H2+H4 was 68.7% under equal priors, 76.0% under the continuity-favouring prior, 84.8% under the literature-neutral prior and 89.3% under the H2-permissive prior. H1 did not lead under any tested family. H3 was competitive only in the equal-prior analysis, which intentionally assigned it a 25% prior despite the absence of BDBV-specific bridge evidence. Prior-family results are summarised in Table 6.

**Table 6.** Model probabilities across prior families and combined support for discontinuous histories.

| Prior family | H1 | H2 | H3 | H4 | H2+H4: discontinuous history |
| --- | --- | --- | --- | --- | --- |
| Equal | 9.6% | 32.6% | 21.6% | 36.2% | 68.7% |
| Continuity-favouring | 19.2% | 36.0% | 4.8% | 40.0% | 76.0% |
| Literature-neutral | 11.1% | 50.0% | 4.2% | 34.7% | 84.8% |
| H2-permissive | 6.8% | 63.6% | 3.8% | 25.7% | 89.3% |

Expressed without Bayesian terminology, every prior family placed most of the weight on histories in which the May epidemic did not descend through one continuously infectious acute chain from January.

The prior sensitivity therefore affected the mechanism selected within the discontinuous class more strongly than it affected the broader conclusion. Across every family, the majority of probability was assigned to histories in which the January signal was not connected to May by one uninterrupted ordinary acute chain.

### 3.3 Posterior timing and transmission requirements

H1 placed the initial seed at median week 2 and the amplification point at week 13. Its median pre-amplification reproduction parameter was 0.98, close to the threshold at which a small overdispersed chain repeatedly risks extinction, and the median post-amplification value was 1.91. H1 therefore required a chain that remained marginal for approximately eleven weeks before accelerating. Selected posterior parameter summaries are reported in Table 7.

**Table 7.** Posterior summaries of selected model parameters.

| Hypothesis | Parameter | Median | 5th percentile | 95th percentile |
| --- | --- | --- | --- | --- |
| H1 | seed_week | 2.00 | 1.00 | 3.00 |
| H1 | amp_week | 13.00 | 11.00 | 14.00 |
| H1 | R_low | 0.98 | 0.61 | 1.23 |
| H1 | R_high | 1.91 | 1.30 | 2.66 |
| H1 | k | 0.24 | 0.05 | 0.75 |
| H2 | early_week | 2.00 | 0.00 | 3.00 |
| H2 | late_week | 11.00 | 9.00 | 13.00 |
| H2 | R_low | 0.85 | 0.49 | 1.20 |
| H2 | R_high | 1.93 | 1.35 | 2.63 |
| H2 | k | 0.25 | 0.07 | 0.72 |
| H3 | early_week | 2.00 | 1.00 | 3.00 |
| H3 | bridge_week | 11.00 | 9.00 | 13.00 |
| H3 | R_low | 0.85 | 0.48 | 1.19 |
| H3 | R_high | 1.96 | 1.38 | 2.72 |
| H3 | k | 0.27 | 0.07 | 0.72 |
| H4 | late_week | 11.00 | 9.00 | 13.00 |
| H4 | R_low | 0.86 | 0.51 | 1.22 |
| H4 | R_high | 1.96 | 1.39 | 2.67 |
| H4 | k | 0.27 | 0.07 | 0.72 |
| H4 | bg_rate | 14.08 | 7.62 | 18.40 |

H2 placed the early event at median week 2 and the later successful introduction at week 11. Its median pre-and post-amplification reproduction parameters were 0.85 and 1.93. H3 placed the survivor-associated bridge at median week 11, making its simulated May histories similar to H2 once reseeding occurred. H4 also placed the independent later seed at median week 11 but required a higher median background event rate than the other models.

### 3.4 Validation, sensitivity and posterior-predictive adequacy

**Figure 5.**
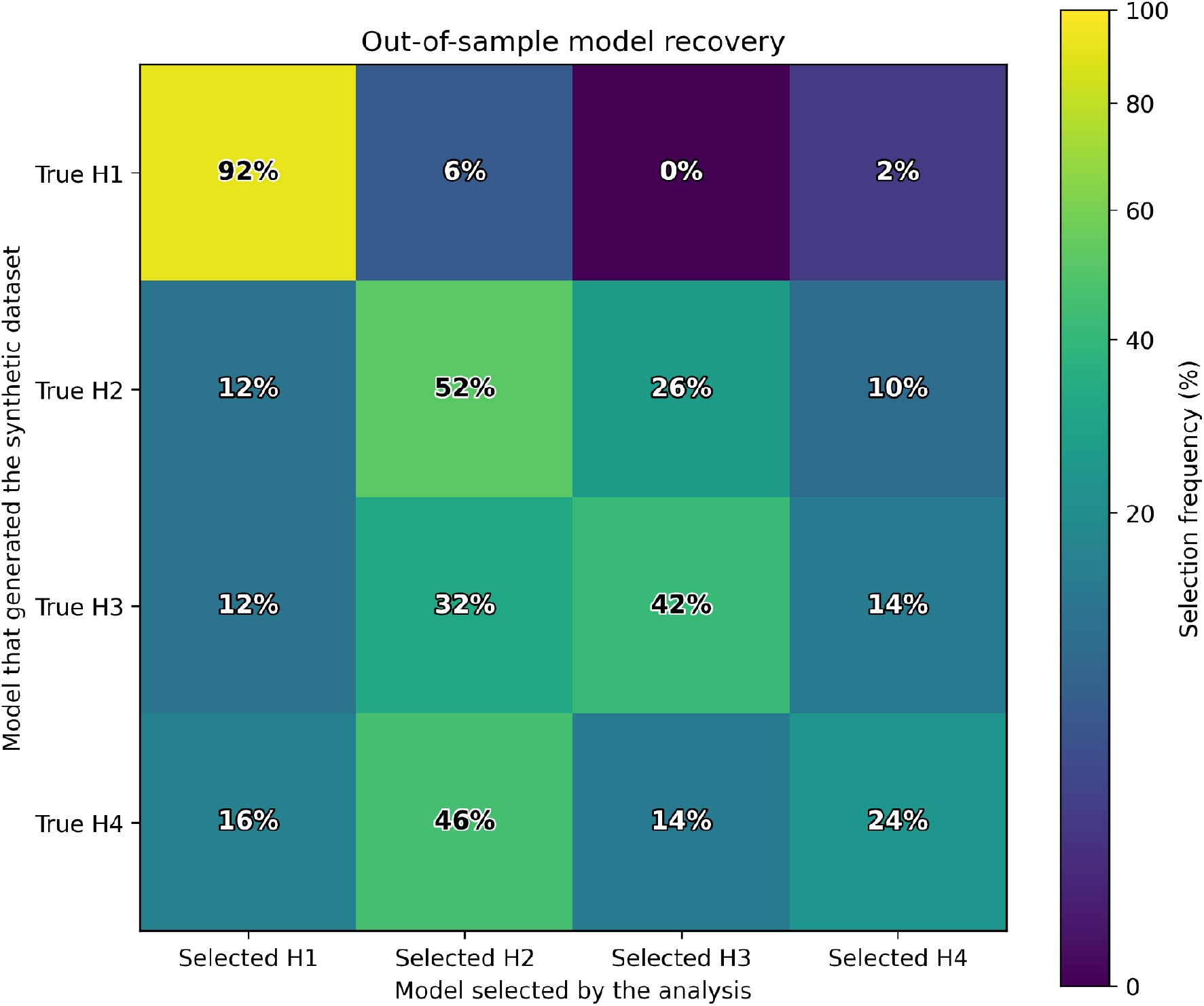
Out-of-sample model recovery under equal priors. Rows denote the model that generated 50 synthetic datasets; columns denote the model selected by the reference-table classifier. Cell values and the colour scale are expressed from 0% to 100%.

H1 was correctly selected in 92% of synthetic datasets, demonstrating that the method could recognise sustained acute continuity when it generated the data. H2 was recovered in 52%, but 26% of H2 datasets were classified as H3. H3 was recovered in 42% and classified as H2 in 32%. H4 was recovered in only 24% and classified as H2 in 46%. The complete recovery matrix is reported in Table 8.

**Table 8.** Out-of-sample model recovery under equal model priors.

| True model | Selected H1 | Selected H2 | Selected H3 | Selected H4 |
| --- | --- | --- | --- | --- |
| H1 | 92% | 6% | 0% | 2% |
| H2 | 12% | 52% | 26% | 10% |
| H3 | 12% | 32% | 42% | 14% |
| H4 | 16% | 46% | 14% | 24% |

A 92% recovery rate means that the analysis recognised 92 of every 100 synthetic H1 outbreaks as H1; the lower diagonal values for H2-H4 show that those discontinuous mechanisms often produce observationally similar evidence.

These results establish an asymmetry in inference. The evidence summaries can identify many continuous-chain histories as H1, but they cannot reliably distinguish whether a later successful seed represents renewed external introduction, survivor reseeding, or an independent outbreak preceded by mixed background mortality. The H2 and H4 probabilities should therefore be interpreted primarily as support for discontinuity, with weaker support for the precise source of that discontinuity.

#### Evidence-component sensitivity

**Figure 6.**
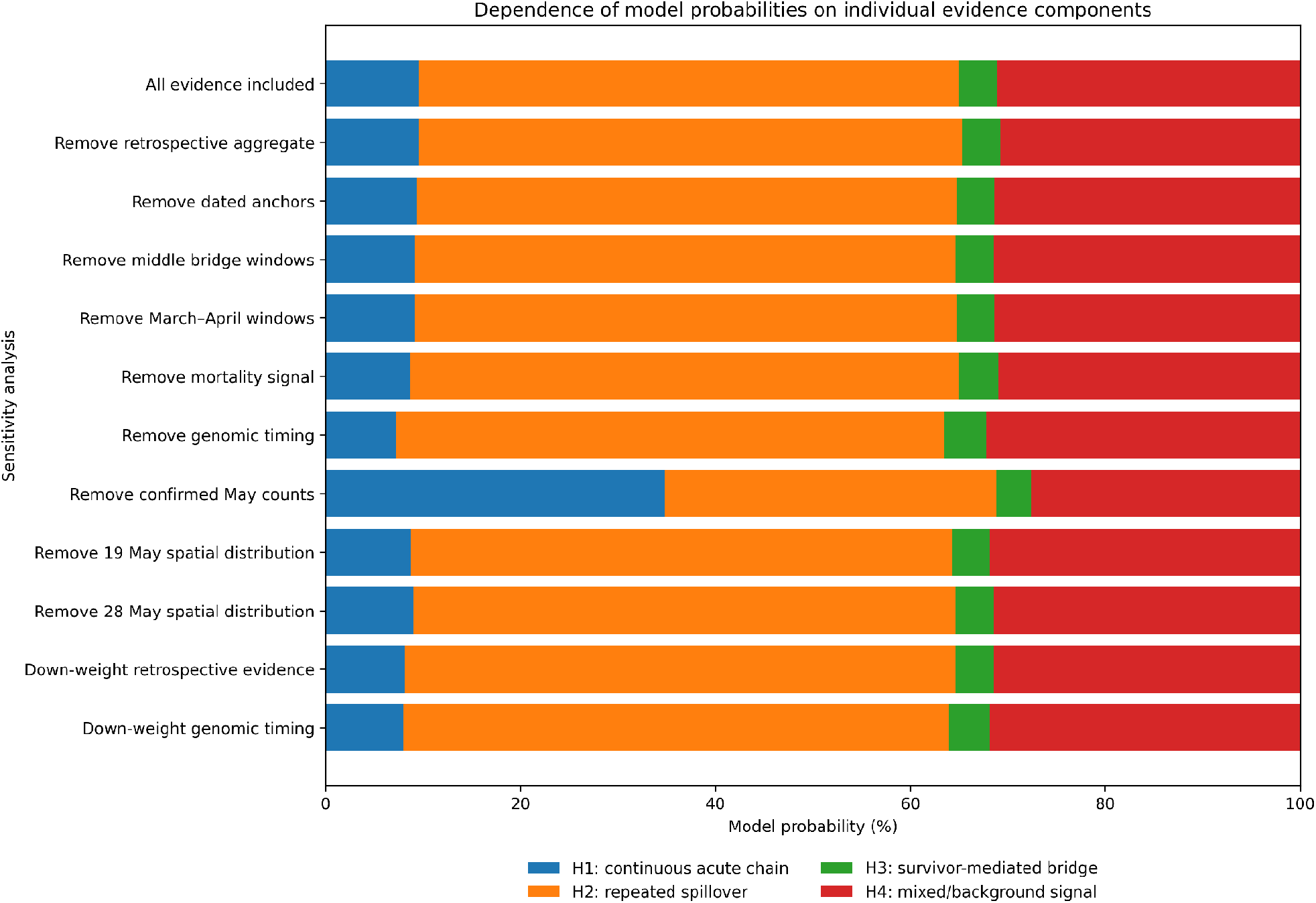
Leave-one-component-out and down-weighting sensitivity under the literature-neutral reference-table analysis. Each bar sums to 100%. The legend is placed below the graph to preserve the full plotting width.

**Figure 7.**
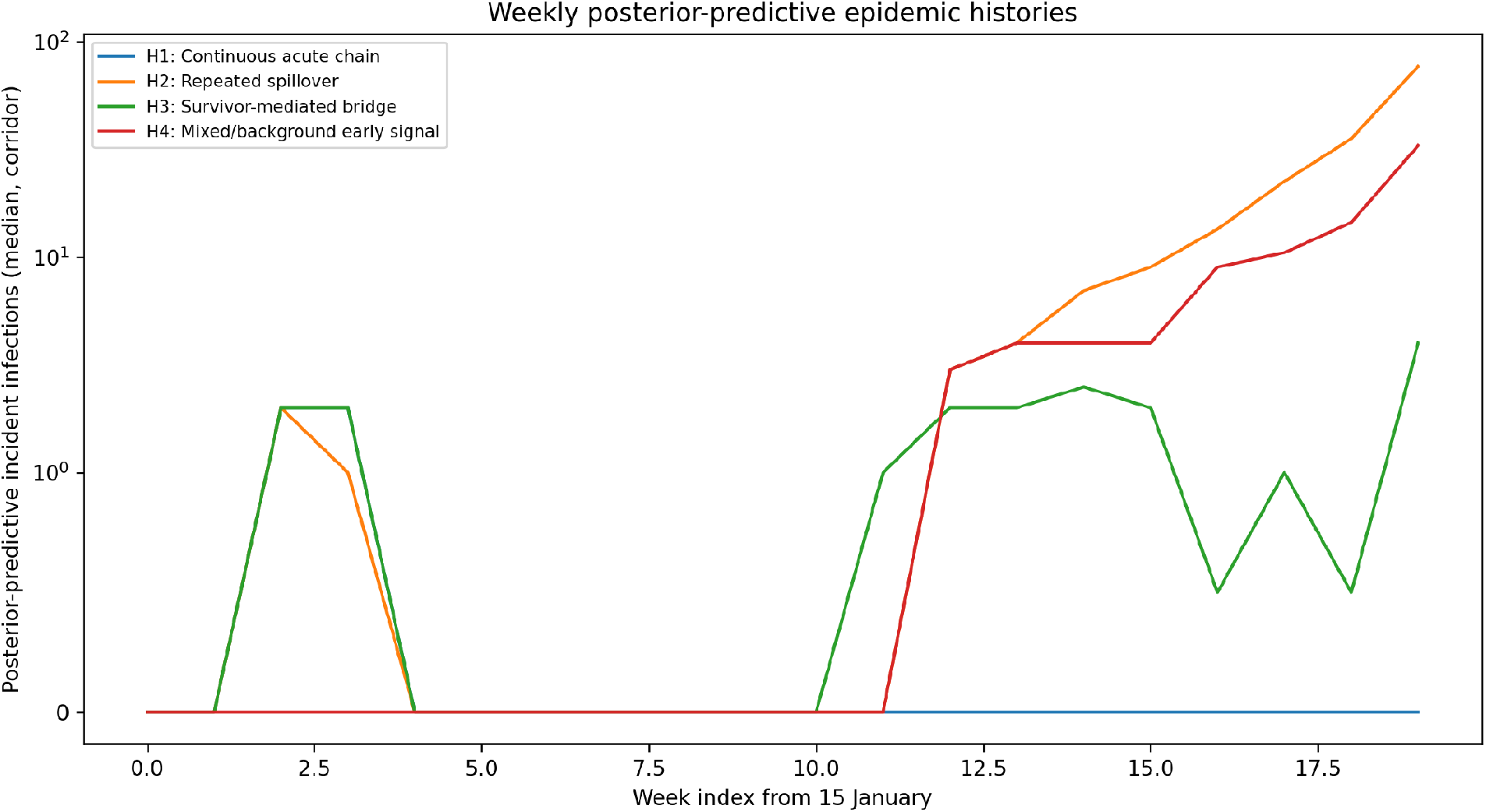
Posterior-predictive incident infections in the four-zone corridor. The trajectories show the weekly pattern each hypothesis required to reproduce the evidence. They are model-generated histories and not estimates of the observed weekly epidemic curve.

With all components included, the reference-table analysis assigned 55.5% to H2, 31.1% to H4, 9.5% to H1 and 3.9% to H3. Removing the retrospective aggregate, dated anchors, bridge-window summaries, mortality signal, genomic timing, or either spatial composition changed individual probabilities by only a few percentage points and did not reverse the H2–H4 ordering.

Removing the confirmed May count trajectory produced the largest change: H1 increased to 34.8%, H2 declined to 34.0%, H4 remained at 27.6%, and H3 remained at 3.6%. The rapid confirmed growth after recognition was therefore the principal evidence distinguishing a later successful seed from a continuously surviving early chain.

Down-weighting the full retrospective bundle did not reduce H2’s support; H2 increased slightly because the May dynamics and later genomic timing were comparatively easier for H2 and H4 to reproduce. This result indicates that the repeated-introduction conclusion was not driven by treating the retrospective aggregate as a confirmed epidemic curve.

#### Posterior-predictive adequacy

Posterior-predictive histories from all four models could reproduce some aspects of the evidence. H1 histories generally required a marginal early chain followed by late amplification. H2 histories contained an early cluster and a later seed near early April. H3 histories resembled H2 after the bridge and were therefore difficult to distinguish without survivor-specific evidence. H4 histories reproduced the recognised outbreak through a later seed while generating more of the January–March signal through the background process.

No single model reproduced every uncertain summary without trade-offs. H1 was penalised by the rapid May trajectory and the need to survive for many weeks. H2 and H4 were penalised by the coherence of the dated early signal, while H3 was penalised by the low prior probability and multiple unobserved bridge requirements. The final probabilities reflect those combined trade-offs rather than one decisive datum.

## 4. DISCUSSION

### 4.1 The robust inference is discontinuity, not proof of one mechanism

The central finding is stronger than a simple ranking of four percentages. Across all tested priors, 68.7%-89.3% of model probability was assigned to H2 or H4, the two histories in which the January/February signal was not connected to the May epidemic by one uninterrupted ordinary acute chain. The combined probability was 84.8% under the main prior. By contrast, H1 and H3 together accounted for only 15.2%. The data therefore favour a later successful origin of the recognised May lineage, while leaving the nature of the earlier Mongbwalu signal only partly identified.

This class-level result is more robust than the distinction between H2 and H4. Equal-prior evidence marginally favoured H4 over H2 by an approximate evidence ratio of only 1.11:1, whereas the literature-neutral prior favoured H2 because direct BDBV precedent justified higher prior probability for multiple introduction. Model recovery further showed that H4 was commonly classified as H2. The scientifically defensible conclusion is therefore that the recognised May lineage most probably arose through a later successful seed or independent onset, while the disease status of the January signal remains incompletely resolved.

In practical epidemiological terms, the January signal should prompt investigation as a real warning event, but it should not be described as the demonstrated direct ancestor of the May epidemic.

### 4.2 Interpretation of the four hypotheses

#### H1 - Continuous acute transmission

H1 cannot be excluded on biological or surveillance grounds. Filovirus outbreaks can remain undetected, early cases can be missed, and sparse genomic sampling can place the common ancestor of sampled viruses later than the first infection. [19,33] The model explicitly allowed an early seed, substantial under-reporting and a later increase in reproduction. It also recovered H1 correctly in 92% of synthetic datasets, indicating that H1’s low probability was not caused by an incapable simulator.

The difficulty was the combination of duration, extinction risk and May growth. The median H1 history required a chain seeded in late January, a pre-amplification reproduction parameter near one, strong overdispersion and an amplification point only in mid-April. Such a chain must remain small enough to avoid recognition, yet avoid extinction through approximately eleven weekly transitions and then expand rapidly enough to match May surveillance. These conditions are possible individually but demanding jointly.

H1 received 9.6% under equal priors and 19.2% even when assigned a 45% continuity-favouring prior. It did not lead under any tested family. We therefore conclude that one uninterrupted ordinary acute chain is not the best-supported explanation of the combined evidence, although an unobserved continuous lineage cannot be ruled out.

#### H2 - Repeated introduction or renewed primary exposure

H2 is supported by a direct species-specific precedent. Additional sequencing of the 2012 Isiro BDBV outbreak produced a topology compatible with more than one spillover and revised the accepted emergence chronology. [1] Concurrent or mixed origins have also been documented in EBOV outbreaks, and clustered primary introductions have occurred during wildlife epizootics and the Durba–Watsa Marburg outbreak. [8,10–13]

The fitted H2 history was epidemiologically coherent. It placed an early event in late January, a later successful seed around early April, subcritical or marginal transmission before amplification, and reproduction close to two afterwards. This structure jointly explains three otherwise difficult features: a structured early Mongbwalu signal, a March-centred sampled ancestor, and explosive confirmed growth in May. It does so without requiring one marginal, overdispersed chain to survive every intervening week. The ablation analysis supports this interpretation: removing the May counts largely removed H2’s advantage, whereas removing genomic timing or the retrospective aggregate did not.

However, H2 is not proven by the posterior percentage. H4 had slightly higher approximate evidence under equal priors, and H2’s 50.0% literature-neutral probability partly reflected the prior probability assigned from the Isiro precedent. Moreover, the model does not identify the source of the later seed.’Repeated spillover’ in this paper includes a renewed primary exposure episode; it should not be interpreted as evidence for a particular animal species, mine exposure or contact route. The BDBV reservoir remains unknown. [7,14–17]

We therefore conclude that H2 is the leading mechanistic explanation after incorporating the relevant BDBV literature, but that the current data establish repeated introduction only as the most plausible specified mechanism, not as a demonstrated event.

#### H3 - Survivor-mediated sexual transmission through persistent virus in semen

H3 operationalised the survivor hypothesis specifically as sexual transmission from a male survivor with persistent virus in semen. It provides a biologically coherent bridge between an early cluster and a later outbreak without requiring continuous acute infectiousness. EBOV RNA can persist in semen for many months, viable virus has been recovered from selected samples, and rare sexual transmission has been supported by epidemiological and genomic linkage. [22–26,28–29] The January-to-early-April interval generated by the model is therefore temporally compatible with the known persistence window.

Temporal compatibility is not equivalent to transmission probability. The semen-mediated pathway requires a sequence of low-frequency events: an infected man must survive the early episode, retain viable rather than merely detectable virus, resume sexual exposure, transmit to a partner, and generate an onward chain that escapes extinction. RNA-positive semen is evidence of persistence, not proof of infectiousness; survivor cohorts have documented prolonged RNA detection and condomless sex without recognised transmission. [27] The probability of the complete bridge is therefore necessarily lower than the probability of RNA persistence alone.

The Mongbwalu evidence did not provide the observations that would specifically favour this pathway. No male survivor from the January/February cluster was linked epidemiologically to a later case, no partner transmission chain was identified, no semen testing result was available, and the genomic evidence did not show the reduced evolutionary accumulation expected after prolonged persistence. In addition, the female-heavy early signal has opposing implications: gendered caregiving can increase female infection risk after household introduction, but it also reduces the expected pool of male survivors capable of semen-mediated transmission. [5,20,30–32]

The model nevertheless allowed a bridge around week 11, approximately early April, and H3 histories could reproduce the later epidemic once reseeding occurred. Their similarity to H2 after reseeding explains why 26% of synthetic H2 datasets were classified as H3 and 32% of H3 datasets as H2. Evidence that would materially increase support for H3 would include a documented male survivor-partner link, positive semen testing near the bridge interval, a compatible contact chain, or genomic evidence of persistence-associated slow evolution.

H3 received 4.2% under the main prior and remained below 5% in all substantive prior families. We therefore conclude that semen-mediated survivor transmission is a credible contingency that should be investigated, but it is not currently supported as the principal origin mechanism.

#### H4 - Mixed or non-ancestral early mortality

H4 is sometimes misread as asserting that the January investigation found nothing. It is broader: the early material may contain unrelated deaths, compatible illness, an extinct BVD cluster, or a mixture of these, while the lineage responsible for the recognised May epidemic begins independently. This formulation reflects the actual evidentiary limitation.

The historical evidence supports caution. During the 2007 BDBV outbreak, a substantial fraction of suspected cases were laboratory negative, and clinical presentation was not sufficiently specific for retrospective diagnosis. [2,4] The Mongbwalu investigation intentionally sought households with clustered deaths and compatible symptoms, so the reported aggregate was signal-enriched rather than a representative diagnostic sample. [21]

H4 had the highest approximate evidence and led under equal and continuity-favouring priors. It was also poorly recoverable: nearly half of synthetic H4 datasets were classified as H2. This shows that a true extinct early cluster and background/mixed mortality can generate similar summary evidence once the May epidemic is introduced later.

We therefore conclude that H4 cannot be rejected and should be presented as a co-leading evidence-based alternative to H2. Additional dated line-list, laboratory or genomic information is required to determine whether the January cluster was BVD, non-BVD, or a mixture.

### 4.3 Triangulation across retrospective, genomic and surveillance evidence

The three evidence layers converge on a limited but important conclusion. The retrospective material supports the existence of an unusual early signal; it does not establish lineage ancestry. The genomic timing supports a later common ancestor for sampled viruses; it does not exclude an earlier extinct or unsampled cluster. The confirmed May trajectory supports a rapidly expanding successful lineage and was the evidence component that most strongly reduced H1.

No individual layer, considered alone, identified H2. Removing genomic timing did not remove H2’s advantage, and removing the retrospective aggregate also did not remove it. Only removal of the May counts eliminated the clear separation between H1 and H2. Conversely, the structured dated early events prevented H4 from becoming overwhelmingly dominant. The model probabilities therefore arose from genuine triangulation: the early signal was too coherent to dismiss completely, the sampled ancestry was too late to establish a direct January lineage, and the May expansion was too rapid to make a long marginal chain the simplest explanation.

This triangulation is also consistent with the recovery results. The data can distinguish many continuous chains from later-onset histories, but cannot reliably determine whether the earlier signal was a genuine extinct cluster or background mortality. The unresolved distinction is therefore empirical, not merely semantic.

### 4.4 Implications, limitations and research priorities

The analysis has implications for how retrospective outbreak investigations are designed and reported. Weekly onset intervals, household and funeral links, healthcare-worker exposures and the distinction between suspected illness and death should be preserved in a shareable analytical dataset. A single cumulative suspected count provides evidence of scale but removes much of the information needed to distinguish continuous transmission, repeated introduction and background mortality.

Genomic sampling should be geographically purposeful. The limited number of Mongbwalu genomes restricts the ability to exclude an early local lineage. During a rapidly expanding outbreak, sequencing only where cases later accumulate can produce a precise description of the dominant lineage while leaving the origin location poorly represented.

Ecological investigation and survivor follow-up should proceed in parallel rather than sequentially. They test different hypotheses and require different evidence. Ecological work should examine shared exposures without assuming a reservoir species; survivor investigation should seek epidemiologically and temporally plausible linkages without treating RNA persistence as proof of infectiousness.

Outbreak communication should distinguish three progressively stronger claims: an early compatible signal existed; compatible events may have continued through the bridge period; and the early signal was directly ancestral to the confirmed outbreak. The present analysis supports the first claim, gives partial support to the second, and does not establish the third.

#### Strengths and limitations

The principal strength of the study is the explicit separation of evidence status. The retrospective aggregate was not treated as a confirmed curve, genomic timing was not treated as an exact origin date, and model probabilities were accompanied by equal-prior evidence, chain stability, recovery and ablation analyses. The weekly spatial simulator also made the distinct requirements of each hypothesis visible rather than embedding them in a qualitative narrative.

The most important limitation is the absence of the underlying retrospective interview database, dated line list and weekly distribution. Consequently, the observation model relied on a published report of the investigation and probability distributions that preserve uncertainty. The January and February events were not laboratory confirmed. A second major limitation is that the genomic component was a soft timing distribution rather than a phylogenetic likelihood based on sequences and tree uncertainty.

The spatial movement matrix was stylised and did not use measured mobility or contact data. The literature review was rapid, unregistered and not independently duplicated. Some evidence summaries originated from the same retrospective material; they were propagated as uncertainty but could not be made fully independent. The model set was also incomplete: multiple introductions combined with short transmission chains, heterogeneous reporting changes and additional survivor or relapse mechanisms were represented only approximately.

Finally, model recovery demonstrated limited identification among H2, H3 and H4. This limitation directly constrains the conclusion. The numerical probabilities should not be presented as proof of one origin event; they quantify how the specified histories compare under the available summaries.

#### Research priorities

The highest-value next step is access to the retrospective investigation’s dated case register or weekly distribution. Incorporating those data would allow explicit transmission-network candidates, separate illness and mortality observation processes, and a better distinction between an extinct early cluster and background mortality.

A second priority is increased genomic representation from Mongbwalu and a formal sequence or tree likelihood. A structured coalescent or simulation-based phylogenetic analysis could directly test whether the sampled March ancestry is compatible with an unsampled January lineage, a later introduction or survivor-associated slowed evolution.

Ecological and occupational exposure histories should be investigated without assuming a bat or mine reservoir, and survivor follow-up records should be reviewed for a plausible male survivor and partner linkage with appropriate confidentiality safeguards. The literature search and bibliography should also be updated immediately before submission.

## 5. CONCLUSION

The hypotheses do not receive equal support. H1, one uninterrupted acute chain from January into May, is possible but is not the leading explanation under any tested prior family; it requires prolonged survival near the extinction threshold followed by sharp amplification. H2, an early cluster followed by a later successful introduction or renewed primary exposure, is the preferred explanation under the literature-informed analysis because it best reconciles the structured early signal, March-centred sampled ancestry and rapid May expansion, and because multiple introduction has direct BDBV precedent. H3, semen-mediated transmission from a male survivor, is temporally and biologically credible but lacks the survivor-partner, semen-testing and genomic evidence needed to make it competitive. H4, a mixed or non-ancestral early signal followed by an independent outbreak, has the strongest equal-prior approximate evidence and remains the principal alternative because the early events were not laboratory confirmed and H4 is frequently confounded with H2.

Our primary conclusion is that the recognised May epidemic most probably did not descend through one continuously transmitting acute chain from January. H2 and H4 together receive 84.8% under the main prior and at least 68.7% under every tested prior family. Within that discontinuous class, H2 is our preferred scientific interpretation: the January/February Mongbwalu signal was likely an epidemiologically meaningful early cluster or warning event, but the lineage that expanded in May most plausibly arose from a later successful introduction or renewed primary exposure. This is not proof of a second zoonotic event, and H4 cannot be excluded. The early signal should therefore be incorporated into the origin investigation without being presented as the demonstrated direct ancestor of the May outbreak. Access to the retrospective line list and more representative early genomic sampling is now the decisive requirement for separating H2 from H4.

## DATA AND CODE AVAILABILITY

The confirmed surveillance data are publicly available from the INRB-UMIE BDBV2026-Data CORE repository (https://github.com/INRB-UMIE/BDBV2026-Data). The analytical code, parameter definitions, evidence distributions, chain diagnostics, recovery outputs, derived tables and figure-generation scripts are supplied with this preprint as supplementary files. The unpublished retrospective line list and interview database were not available to the authors.

## ETHICS STATEMENT

This study used only publicly available, aggregated and non-identifiable secondary data and did not involve recruitment, interaction with human participants, or access to private identifiable information. Ethics committee approval and informed consent were therefore not applicable to this modelling analysis.

## GENERATIVE AI STATEMENT

Generative artificial intelligence was used under author supervision to assist literature organisation, code development, document preparation and language editing. The authors are responsible for verification of all evidence classifications, model definitions, calculations, citations and interpretations.

## AUTHOR CONTRIBUTIONS

Johan G.L. Verheyden: Conceptualisation; Methodology; Software; Validation; Formal analysis; Investigation; Resources; Data curation; Visualisation; Project administration; Writing - original draft. Celestin Nzanzu Mudogo: Writing - review and editing; final substantive review. Wolfgang Jacquet: Writing - review and editing; final substantive review.

## FUNDING

This work received no specific funding.

## COMPETING INTERESTS

The authors declare no competing interests.

## SUPPLEMENTARY MATERIAL

Supplementary Methods, Supplementary Figure S1, and Supplementary Tables S1-S8 are provided in the accompanying supplementary-material file. Full machine-readable tables, analytical outputs, Python and R scripts, and reproduction instructions are provided in the accompanying Data and Code archive.

## Data Availability

All data produced are available online at

https://github.com/INRB-UMIE/BDBV2026-Data

## Notes

### Competing Interest Statement

The authors have declared no competing interest.

### Author Declarations

All confirmed surveillance data used in this study were obtained from publicly available, aggregated sources. National and health-zone surveillance data are available from the INRB-UMIE BDBV2026-Data CORE repository. The analytical code, parameter definitions, evidence distributions, chain diagnostics, model-recovery outputs, derived tables and figure-generation scripts are included with this preprint as supplementary files. The unpublished retrospective line list and interview database described in secondary reporting were not available to the authors and are therefore not included in the shared data.

