## Supplement for "ONE CHAIN, REPEATED SPILLOVER, OR AN UNRELATED EARLY SIGNAL?"

### SUPPLEMENTARY METHODS S1. EVIDENCE REPRESENTATION

The analysis separated latent outbreak histories from observation processes. Retrospective evidence was represented as uncertain compatibility evidence rather than a confirmed epidemic curve. The evidence kernel was integrated across 48 draws from the uncertainty distributions. Official confirmed counts and early health-zone composition constrained the May epidemic, while genomic timing was entered as a soft sampled-ancestry constraint rather than a sequence likelihood.

| Component | Observed basis | Model distribution | Interpretation boundary |
|---|---|---|---|
| Retrospective aggregate | 500 reports, conservative lower bound | Beta-binomial via q~Beta(2,18) | Secondary/unpublished underlying investigation; not a confirmed curve |
| Dated anchors | Six household/funeral events | Beta-binomial via q_anchor~Beta(3,12) | Dated but unconfirmed |
| Middle bridge | Four 21-day middle windows | Binomial(4,0.8152) | Propagated from v0.2; not independent raw evidence |
| March/early April | Two bridge windows | Binomial(2,0.8753) | Propagated from v0.2; not independent raw evidence |
| Mortality signal | Up to 108 reported deaths | Beta-binomial via q_death~Beta(2,18) | Upper-bound retrospective mortality signal |
| Genomic timing | Early/mid-March centre; Feb-Apr interval | Truncated Normal(centre day 55, SD 22, day 17-87) | Soft combined constraint, not a sequence likelihood |
| Confirmed counts | 8, 51 and 210 on 14, 19 and 28 May | Negative-binomial observation distributions | Official CORE surveillance data |
| Spatial composition | Mongbwalu/Bunia/Rwampara/ Nizi on 19 and 28 May | Dirichlet distributions around official proportions | Allows reporting revisions and incomplete early reconciliation |

### SUPPLEMENTARY METHODS S2. WEEKLY SPATIAL SIMULATOR

The simulator covered 20 weekly periods beginning 15 January 2026 and four health zones: Mongbwalu, Bunia, Rwampara and Nizi. Weekly infections followed an overdispersed negative-binomial branching process. A movement parameter distributed a proportion of infections across the four-zone corridor. H1 contained one early introduction and a later amplification point; H2 contained early and later introductions; H3 contained an early introduction and survivor-associated reseeding; H4 contained an independent later introduction and a separate background-event process.

The retrospective observation process was $Y(t,z) = \text{Binomial}[I(t,z), q_{case}] + \text{Binomial}[B(t,z), q_{bg}]$. The q parameters represent inclusion in a compatibility curve, not diagnostic probability. Prospective confirmed reports were generated through an uncertain reporting fraction.

### SUPPLEMENTARY METHODS S3. ABC-SMC IMPLEMENTATION

For each hypothesis, three independent chains of 450 particles were run over tolerances 3.0, 2.4, 1.9, 1.5, 1.25, 1.05 and 0.90. At each stage particles were reweighted with a Gaussian ABC kernel, systematically resampled, and rejuvenated through reflected random-walk Metropolis proposals. Approximate model evidence was accumulated sequentially. Four prior families were retained to display rather than conceal prior sensitivity.

| Prior family | H1 | H2 | H3 | H4 |
|---|---|---|---|---|
| Equal | 0.25 | 0.25 | 0.25 | 0.25 |
| Continuity-favouring | 0.45 | 0.25 | 0.05 | 0.25 |
| Literature-neutral | 0.30 | 0.40 | 0.05 | 0.25 |
| H2-permissive | 0.20 | 0.55 | 0.05 | 0.20 |

### SUPPLEMENTARY METHODS S4. VALIDATION AND SENSITIVITY

Model recovery used 50 independent synthetic datasets per generating model and a separate reference table of 800 simulations per model. The recovery matrix was used to evaluate identifiability, not to mechanically correct model probabilities. Leave-one-component-out analyses removed each evidence layer in turn, and additional analyses down-weighted the retrospective bundle or genomic timing.

### SUPPLEMENTARY TABLES

The complete supplementary tables are provided as machine-readable CSV files. Selected compact tables are reproduced below; large matrices and chain-level outputs are supplied in full in the accompanying files.

#### Supplementary Table S2

Supplementary Table S2. Evidence provenance and uncertainty definitions.

| component | observed_basis | model_distribution | interpretation_boundary |
|---|---|---|---|
| Retrospective aggregate | 500 reports, conservative lower bound | Beta-binomial via q~Beta(2,18) | Secondary/unpublished underlying investigation; not a confirmed curve |
| Dated anchors | Six household/funeral events | Beta-binomial via q_anchor~Beta(3,12) | Dated but unconfirmed |
| Middle bridge | Four 21-day middle windows | Binomial(4,0.8152) | Propagated from v0.2; not independent raw evidence |
| March/early April | Two bridge windows | Binomial(2,0.8753) | Propagated from v0.2; not independent raw evidence |
| Mortality signal | Up to 108 reported deaths | Beta-binomial via q_death~Beta(2,18) | Upper-bound retrospective mortality signal |
| Genomic timing | Early/mid-March centre; Feb-Apr interval | Truncated Normal(centre day 55, SD 22, day 17-87) | Soft combined constraint, not a sequence likelihood |
| Confirmed counts | 8, 51 and 210 on 14, 19 and 28 May | Negative-binomial observation distributions | Official CORE surveillance data |
| Spatial composition | Mongbwalu/Bunia/Rwampara/Nizi on 19 and 28 May | Dirichlet distributions around official proportions | Allows reporting revisions and incomplete early reconciliation |

### Supplementary Table S8

Supplementary Table S8. Combined approximate model evidence.

| hypothesis | combined_logZ | chain_mean | chain_sd | chain_min | chain_max |
|---|---|---|---|---|---|
| H1 | -4.486751531754117 | -4.490080411323805 | 0.1009477460716213 | -4.606493136747395 | -4.426725703583804 |
| H2 | -3.2680019099173494 | -3.2695841837258226 | 0.06935641688716321 | -3.3493119484254654 | -3.2231685676035724 |
| H3 | -3.6760167429696002 | -3.677669816228113 | 0.07091001979518641 | -3.7593328718974632 | -3.631681747749541 |
| H4 | -3.16233971425918 | -3.1624623322903322 | 0.01919703220759415 | -3.1832492171022944 | -3.1454014233160006 |

### Supplementary Table S5

Supplementary Table S5. Model-recovery results.

| recovery_prior | true_hypothesis | selected_hypothesis | selection_frequency | mean_assigned_probability | n_synthetic_datasets |
|---|---|---|---|---|---|
| equal | H1 | H1 | 0.92 | 0.4239689048178688 | 50 |
| equal | H1 | H2 | 0.06 | 0.18205827598250846 | 50 |
| equal | H1 | H3 | 0.0 | 0.1900563964930516 | 50 |
| equal | H1 | H4 | 0.02 | 0.20391642270657123 | 50 |
| literature_neutral | H1 | H1 | 0.9 | 0.48862155279809466 | 50 |
| literature_neutral | H1 | H2 | 0.1 | 0.27886756387561534 | 50 |
| literature_neutral | H1 | H3 | 0.0 | 0.036546039708054784 | 50 |
| literature_neutral | H1 | H4 | 0.0 | 0.19596484361823527 | 50 |
| equal | H2 | H1 | 0.12 | 0.14884310904375306 | 50 |
| equal | H2 | H2 | 0.52 | 0.3025706622677153 | 50 |
| equal | H2 | H3 | 0.26 | 0.2754284634137131 | 50 |
| equal | H2 | H4 | 0.1 | 0.2731577652748187 | 50 |
| literature_neutral | H2 | H1 | 0.12 | 0.18332775397699558 | 50 |
| literature_neutral | H2 | H2 | 0.86 | 0.4856979991964853 | 50 |
| literature_neutral | H2 | H3 | 0.0 | 0.05578740355067436 | 50 |
| literature_neutral | H2 | H4 | 0.02 | 0.27518684327584464 | 50 |
| equal | H3 | H1 | 0.12 | 0.17259344168646368 | 50 |
| equal | H3 | H2 | 0.32 | 0.275782508577499 | 50 |
| equal | H3 | H3 | 0.42 | 0.2839467151276894 | 50 |
| equal | H3 | H4 | 0.14 | 0.2676773346083479 | 50 |
| literature_neutral | H3 | H1 | 0.1 | 0.21636110861973942 | 50 |
| literature_neutral | H3 | H2 | 0.82 | 0.45026974149064025 | 50 |
| literature_neutral | H3 | H3 | 0.0 | 0.0585446000909298 | 50 |
| literature_neutral | H3 | H4 | 0.08 | 0.27482454979869053 | 50 |
| equal | H4 | H1 | 0.16 | 0.147212526445927 | 50 |
| equal | H4 | H2 | 0.46 | 0.2930042119796514 | 50 |
| equal | H4 | H3 | 0.14 | 0.26780154024469693 | 50 |
| equal | H4 | H4 | 0.24 | 0.29198172132972466 | 50 |
| literature_neutral | H4 | H1 | 0.16 | 0.1818509682803336 | 50 |
| literature_neutral | H4 | H2 | 0.78 | 0.4695187340691211 | 50 |
| literature_neutral | H4 | H3 | 0.0 | 0.054206030603384185 | 50 |
| literature_neutral | H4 | H4 | 0.06 | 0.2944242670471612 | 50 |

### Supplementary Table S6

Supplementary Table S6. Evidence-ablation and down-weighting results.

| scenario | hypothesis | hypothesis_name | probability |
|---|---|---|---|
| none | H1 | Continuous acute chain | 0.09539353898556911 |
| none | H2 | Repeated spillover | 0.5545451872609847 |
| none | H3 | Survivor-mediated bridge | 0.03927726037112115 |
| none | H4 | Mixed/background early signal | 0.310784013382325 |
| omit_retrospective_total | H1 | Continuous acute chain | 0.09525566978176159 |
| omit_retrospective_total | H2 | Repeated spillover | 0.5582204657899811 |
| omit_retrospective_total | H3 | Survivor-mediated bridge | 0.03948144148328441 |
| omit_retrospective_total | H4 | Mixed/background early signal | 0.307042422944973 |
| omit_anchors | H1 | Continuous acute chain | 0.09298254075632735 |
| omit_anchors | H2 | Repeated spillover | 0.5545969577019538 |
| omit_anchors | H3 | Survivor-mediated bridge | 0.03864824438261873 |
| omit_anchors | H4 | Mixed/background early signal | 0.3137722571591002 |
| omit_middle_bridge | H1 | Continuous acute chain | 0.0916023264691415 |
| omit_middle_bridge | H2 | Repeated spillover | 0.5550978487906487 |
| omit_middle_bridge | H3 | Survivor-mediated bridge | 0.03894084147501316 |
| omit_middle_bridge | H4 | Mixed/background early signal | 0.31435898326519646 |
| omit_march_april | H1 | Continuous acute chain | 0.09111657566021891 |
| omit_march_april | H2 | Repeated spillover | 0.5564023094628134 |
| omit_march_april | H3 | Survivor-mediated bridge | 0.038915416297440955 |
| omit_march_april | H4 | Mixed/background early signal | 0.3135656985795268 |
| omit_deaths | H1 | Continuous acute chain | 0.0863703459227536 |
| omit_deaths | H2 | Repeated spillover | 0.563565818214417 |
| omit_deaths | H3 | Survivor-mediated bridge | 0.03999308584892002 |
| omit_deaths | H4 | Mixed/background early signal | 0.31007075001390944 |
| omit_genomics | H1 | Continuous acute chain | 0.07277643131550432 |
| omit_genomics | H2 | Repeated spillover | 0.5620562261660295 |
| omit_genomics | H3 | Survivor-mediated bridge | 0.04338800415505084 |
| omit_genomics | H4 | Mixed/background early signal | 0.32177933836341566 |
| omit_may_counts | H1 | Continuous acute chain | 0.3477806240758915 |
| omit_may_counts | H2 | Repeated spillover | 0.340158796034536 |
| omit_may_counts | H3 | Survivor-mediated bridge | 0.036317018037052666 |
| omit_may_counts | H4 | Mixed/background early signal | 0.27574356185251986 |
| omit_spatial_may19 | H1 | Continuous acute chain | 0.08731563871159738 |
| omit_spatial_may19 | H2 | Repeated spillover | 0.555721640232034 |
| omit_spatial_may19 | H3 | Survivor-mediated bridge | 0.03856524565124059 |
| omit_spatial_may19 | H4 | Mixed/background early signal | 0.31839747540512825 |
| omit_spatial_may28 | H1 | Continuous acute chain | 0.08943521138199154 |
| omit_spatial_may28 | H2 | Repeated spillover | 0.557227996918549 |
| omit_spatial_may28 | H3 | Survivor-mediated bridge | 0.03828065885100165 |
| omit_spatial_may28 | H4 | Mixed/background early signal | 0.31505613284845807 |

| scenario | hypothesis | hypothesis_name | probability |
|---|---|---|---|
| downweight_retrospective_bundle | H1 | Continuous acute chain | 0.08099273663859301 |
| downweight_retrospective_bundle | H2 | Repeated spillover | 0.5653704902801792 |
| downweight_retrospective_bundle | H3 | Survivor-mediated bridge | 0.03887145950804707 |
| downweight_retrospective_bundle | H4 | Mixed/background early signal | 0.31476531357318077 |
| downweight_genomics | H1 | Continuous acute chain | 0.07971069968250694 |
| downweight_genomics | H2 | Repeated spillover | 0.5599999723145804 |
| downweight_genomics | H3 | Survivor-mediated bridge | 0.04199336950512772 |
| downweight_genomics | H4 | Mixed/background early signal | 0.3182959584977847 |

### SUPPLEMENTARY FIGURE S1

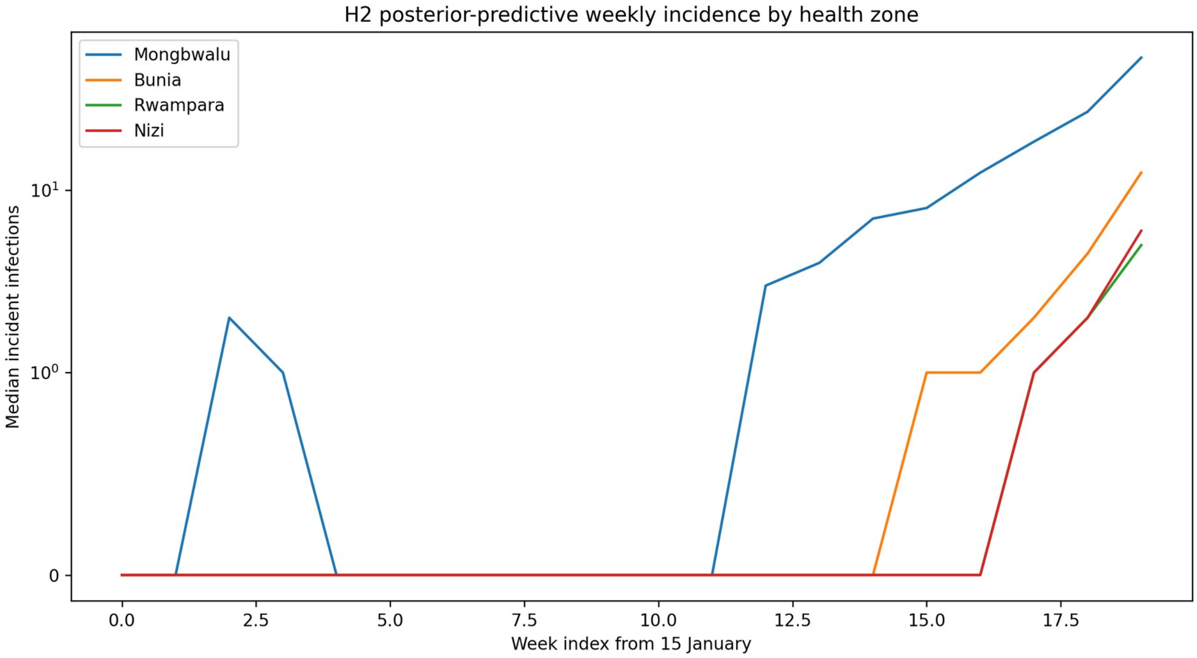


**Supplementary Figure S1. H2 posterior-predictive weekly incidence by health zone.**

### REPRODUCIBILITY

The accompanying Data and Code archive contains the Python and R scripts, input evidence tables, complete derived outputs, fixed random seed, configuration file and checksums needed to reproduce the reported tables and figures. The simulator remains an explicit model of weekly histories and is not a patient-level transmission reconstruction or a phylogenetic sequence likelihood.
